# Breaking New Ground in Computational Psychiatry: Model-Based Characterization of Forgetting in Healthy Aging and Mild Cognitive Impairment

**DOI:** 10.1101/2023.05.13.23289941

**Authors:** Holly Sue Hake, Bridget Leonard, Sara Ulibarri, Thomas Grabowski, Hedderik Van Rijn, Andrea Stocco

**Affiliations:** Department of Psychology, University of Washington, Seattle, WA 98195 USA; Department of Radiology, University of Washington, Seattle, WA 98195 USA; Department of Experimental Psychology, University of Groningen, Groningen, 9712 CP Netherlands

**Keywords:** memory, forgetting, dementia, ACT-R

## Abstract

Computational models of memory used in adaptive learning settings trace a learner’s memory capacities. However, less work has been done on the implementation of these models in the clinical realm. Current assessment tools lack the reliable, convenient, and repeatable qualities needed to capture the individualized and evolving nature of memory decline. The goal of this project was to predict and track memory decline in subjectively- or mildly cognitively impaired (MCI) individuals by using a model-based, adaptive fact-learning system. Here we present data demonstrating that these tools can diagnose mild memory impairment with over 80% accuracy after a single 8-minute learning session. These findings provide new insights into the nature and progression of memory decline and may have implications for the early detection and management of Alzheimer’s disease and other forms of dementia.

## Introduction

Memory and cognitive impairments are a common and debilitating aspect of aging, particularly in conditions such as dementia due to Alzheimer’s Disease (AD). Despite significant efforts to understand and treat these conditions, progress has been slow. One major challenge is the lack of understanding of the relationship between long-term memory decline and the underlying neuropathology. To gain a better understanding of this relationship, it is necessary to have precise and longitudinal assessments of memory function that can identify which aspect of long-term memory is declining, at what rate, and due to which underlying affected neural circuit. However, current clinical tools for memory assessments are not adequate for this purpose. They are often only administered by experts, cannot be performed frequently, lack transparency in interpretation, and are not specific (e.g., not distinguishing between true forgetting and retrieval difficulties, especially in mildly affected patients).

One potential solution to understanding the relationship between long-term memory decline and neuropathology is the use of computational cognitive models. Here, we employed a cognitive model that simulates encoding and passive forgetting based on established cognitive and biological principles, providing a framework for understanding the underlying mechanisms of memory decline in aging and neurodegenerative conditions. This approach can be used to generate predictions about the progression of memory decline and identify potential therapeutic targets.

## Model

The model used herein was originally developed by Anderson & Schooler (1991) within the ACT-R architecture. Consistent with Multiple Trace Theory (Nadel et al., 2000), the model assumes that a memory is made of individual traces created every time the same information is encountered. Each trace decays according to the power law of forgetting (Newell & Rosenbloom, 1982). The odds of retrieving a memory *m* at time *t* are proportional to its activation *A*(*m, t*), which represents the log odds of retrieving any of its component traces, as shown in Eq. (1)

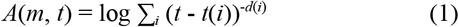

Where *t*(*i*) is the creation time of the *i*-th trace, and *d*(*i*) is its characteristic power decay rate. This trace-specific decay rate, in turn, depends on the residual activation of the memory at the time the trace was created (Pavlik & Anderson, 2005; Sense et al., 2016):

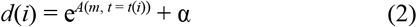

Because Eq. (2) makes the decay rate of each trace dependent on the memory’s activation, it provides an explanation for the spacing effect (Cepeda et al., 2008). That is, traces closer in time have higher decay rates because of the greater activation *A*(*m, t*) of the memory at time *t*(*i*).

Note that this model still depends on a single parameter, α, which we refer to as the *speed of forgetting (SoF)*. The *speed of forgetting* explains the relationship between the history of a memory and the likelihood of being able to retrieve it in the future. Thus, the odds of being able to recall a memory at a later time depend solely on the rate at which the memory is forgotten. Additionally, this suggests that by looking at the history of a memory and the number of times it has been assessed, it is possible to determine the rate at which that memory is forgotten.

This model has been previously used as a cognitive support tool to optimize student learning (Sense & Rijn, 2022; Sense, Velde, & Rijn, 2021; van Rijn et al., 2009; Wilschut, van der Velde, Sense, Fountas, & van Rijn, 2021). In this paradigm, students use the software to learn new facts. The software uses the students’ responses to estimate the *speed of forgetting* value for each fact and optimizes the time and frequency of presentation to maximize the number of facts being memorized. Van Rijn et al. (2009) greatly improved on this design by using the students’ response times, in addition to errors, to estimate *speed of forgetting*.

Sense et al found that *speed of forgetting* was characteristic of an individual, and was highly stable (*r* > 0.6) across time and materials (2016). Furthermore, using neuroimaging methods, Zhou et al. (2021) and Xu et al. (2021) found that not only does *speed of forgetting* capture individual differences in long-term memory function, but also correlates with, and can be decoded from, their spontaneous brain activity at rest.

## Experimental Hypotheses

Based on the behavioral and imaging findings, we hypothesized that the *speed of forgetting* successfully summarizes, at a computational level, the different biological processes of passive forgetting (Davis & Zhong, 2017). These processes include loss of context clues, retrieval interference from other similar memories, and “natural” biological decay. Critically, some of these processes are accelerated in aging (Shuai et al., 2010) and abnormally elevated in amnestic dementias, such as AD. Thus, the model outlined above should be able to distinguish between abnormal memory impairments and normal aging controls. If successful, this model could become an important tool in the clinical assessment of memory function. Furthermore, if the model-based assessment is robust to practice effects and can be repeated frequently, it could provide a new, highly detailed view of memory decay trajectories in normal and abnormal aging, and of the effects of interventions.

To test these ideas, we conducted a longitudinal study of healthy elderly adults and elderly individuals with Mild Cognitive Impairment (MCI). We chose individuals with MCI, rather than dementia, to both ensure participants had sufficient cognitive abilities to perform the task and test the model’s ability to detect earlier, more subtle differences in memory function as it is often a precursor to AD and other forms of dementia (Petersen et al., 2009). This cohort of individuals was followed for 6+ months, during which they performed weekly online model-based assessments to characterize their *speed of forgetting*. We hypothesized that (1) individuals with MCI would exhibit larger *SoF* than healthy controls; (2) *SoF* values would be reliable across repeated assessments; (3) *SoF* values would have clinical validity; i.e. it would be possible to identify differences in abnormal memory function from an individual *SoF*; and finally (4) *SoF* would increase over a period of months, capturing the trajectory of abnormal and healthy aging.

## Materials and Methods

### Participants

Sixteen participants were recruited on a rolling basis from the local NIH-designated Alzheimer’s Disease Research Center. The inclusion criteria for the study were as follows: (1) age between 55 and 85 years, (2) fluency in English, and (3) no major medical or psychiatric conditions that would affect cognitive performance. Participants were classified into two groups: healthy cognition (N = 7; 3F aged 58-71, 4M aged 57-71) and those with mild cognitive impairment (MCI; N = 9; 2F aged 63, 7M aged 67-78).

#### Mild Cognitive Impairment

MCI can be defined as a decline in cognitive abilities that is greater than what is typical for a person’s age and educational background but does not meet the criteria for a diagnosis of dementia (Winblad et al., 2004). MCI was diagnosed using a combination of methods including clinical evaluation, cognitive testing, and medical history. The clinical evaluation was conducted by a geriatric psychiatrist or a neurologist, who assessed the participant’s cognitive and functional abilities using standardized tools. Cognitive testing was performed using a battery of neuropsychological tests that measured various cognitive domains such as memory, attention, and executive function. Medical history was obtained through a structured interview and review of medical records. Participants were classified as having MCI if they had a Clinical Dementia Rating scale <= 0.5.

Additionally, individuals with *subjective* reports of decline by self and/or informant in conjunction with objective cognitive deficits were also included in the MCI group. Healthy controls were screened for cognitive impairment using the same methods as MCI participants. They were classified as healthy controls if they scored within normal limits on cognitive tests and had no history of cognitive decline or functional impairment. All participants provided informed consent and were compensated for their participation in the online memory game portion of the study. All recruitment and testing procedures were approved by the University’s Institutional Review Board.

### Adaptive Memory Assessment

Weekly at-home assessments were completed with the online adaptive fact learning system (AFLS) described in Sense et al. (2016). This system continuously estimates the individualized *speed of forgetting* values in real time as the participant works through the lesson. The software was designed so that participants could perform the task from home using any mobile device. The AFLS works by presenting new study pairs (e.g., “France / Paris”) and scheduling repeated tests (e.g., “France / ?”) at strategic points based on the online estimates of a user’s *speed of forgetting*. Figure 1 provides an example of the software interface.

**Figure 1.**
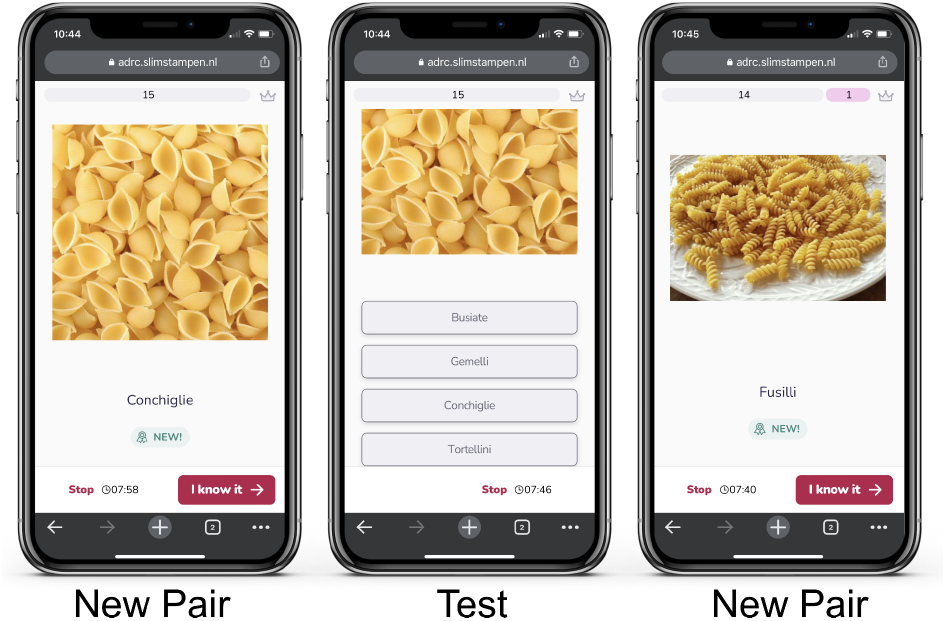
Interface of adaptive fact learning software on a mobile phone.

#### Study Materials

Thirty-two lessons were prepared in advance, spanning different topics (such as European capitals, Swahili words, Asian flags, bird species, types of pasta, flower species). The materials were vetted prior to the experiment to make sure they were comparable in terms of familiarity and difficulty. For each lesson, 15 different pairs were created, each of which associated an object with an English noun. In half of the pairs, the object was presented as an image (e.g., a picture of a Starling with the name “Starling” for the Bird’s lesson), and in the other half, the object was a word (e.g., “France” / “Paris” for European capitals). This was done to investigate possible differences due to the encoding modality (verbal vs. visual objects). The number of terms reached in each lesson depended on the response times and errors of the individual.

#### Data Processing

The repetition, activation, and *speed of forgetting* values for each term were calculated using functions from the software package. The average *speed of forgetting* values for each lesson and the individual were identified by using the terminal α value of each pair at the very last repetition of that term. The data was then filtered to only contain the first full session of a topic (>6 min). This was needed to eliminate any superfluous sessions (some participants desired to complete the task more than once). The data was also organized by the week the lesson was completed to view temporal trends.

## Results

Figure 2 provides an overview of the results. Across all the topics and materials, individual *speeds of forgetting* varied between 0.29 and 0.58 and were normally distributed. *Speed of forgetting* across lessons ranged from *α* = 0.29 to *α* = 0.55, with a mean of *α* = 0.4. The results produced in the present study reproduced the main finding in Sense et al. (2016), wherein individuals’ *speeds of forgetting* were generally stable over time, but differed slightly across materials.

**Figure 2.**
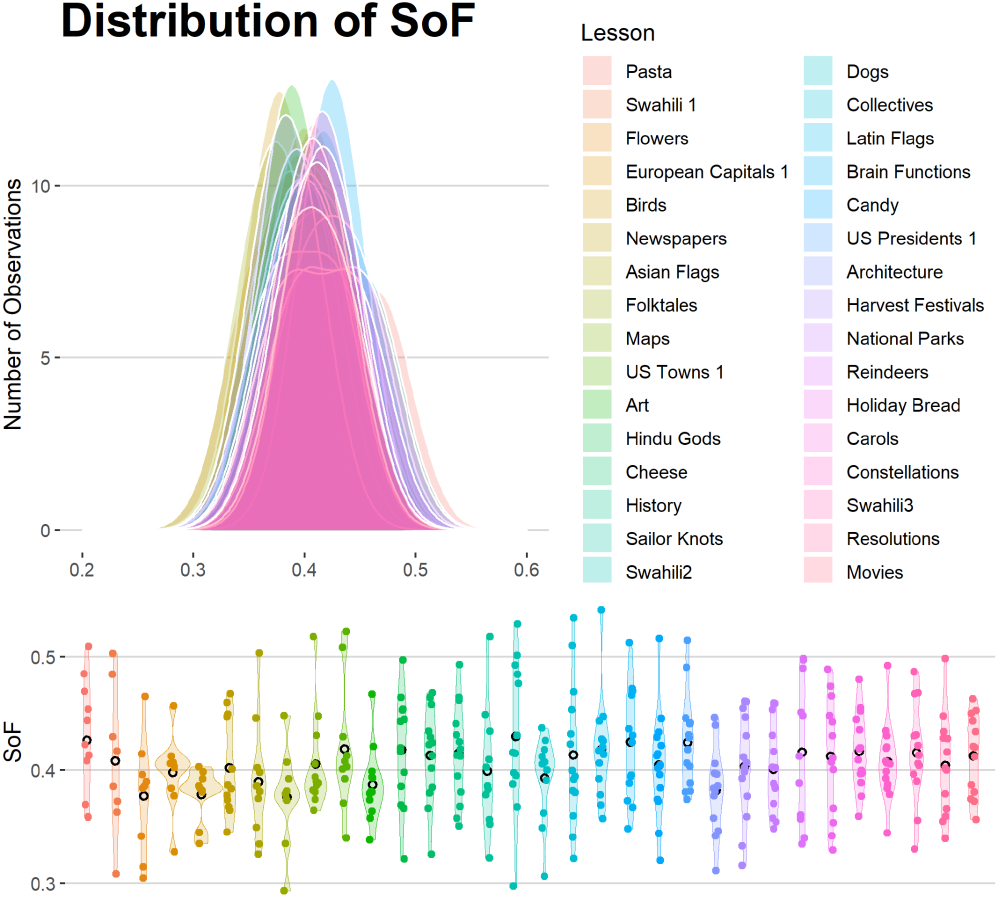
Distribution of *speed of forgetting* across lessons.

### Reliability of the *Speed of Forgetting*

The test-retest reliability of the *speed of forgetting* parameter across materials was assessed using pairwise Pearson correlations between every pair of lessons. The parameter was found to have a high average correlation coefficient of *r* = 0.70 (Fig 3).

**Figure 3.**
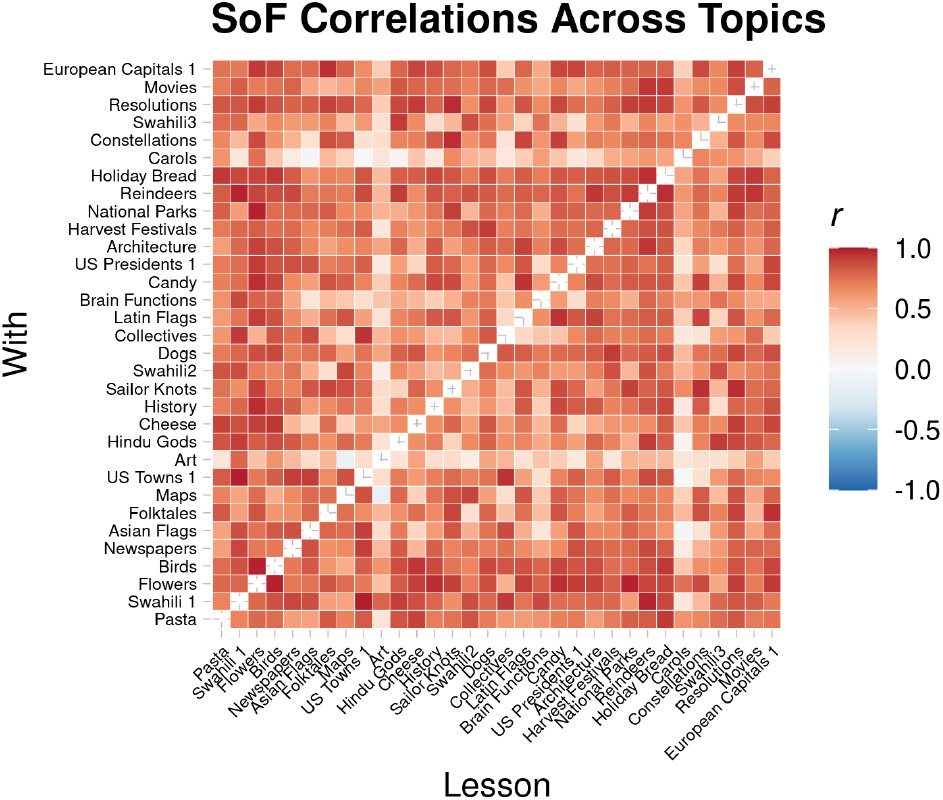
Test-retest reliability of *speed of forgetting*.

### Global Differences Between MCI and Controls

Differences in *speed of forgetting* for healthy controls and individuals diagnosed with MCI as per the gold standard clinical assessment were compared. On average, healthy controls had a *speed of forgetting* of *α* = 0.39, whereas MCI had a *speed of forgetting* of *α* = 0.42 (Fig 4). The difference between groups was compared using a mixed-effects linear model that included the specific weekly topic as a random effect (to account for differences in familiarity). The linear model confirmed the existence of a large main effect of group (β = 0.04, *t* = 9.78, *p* < 0.0001).

**Figure 4.**
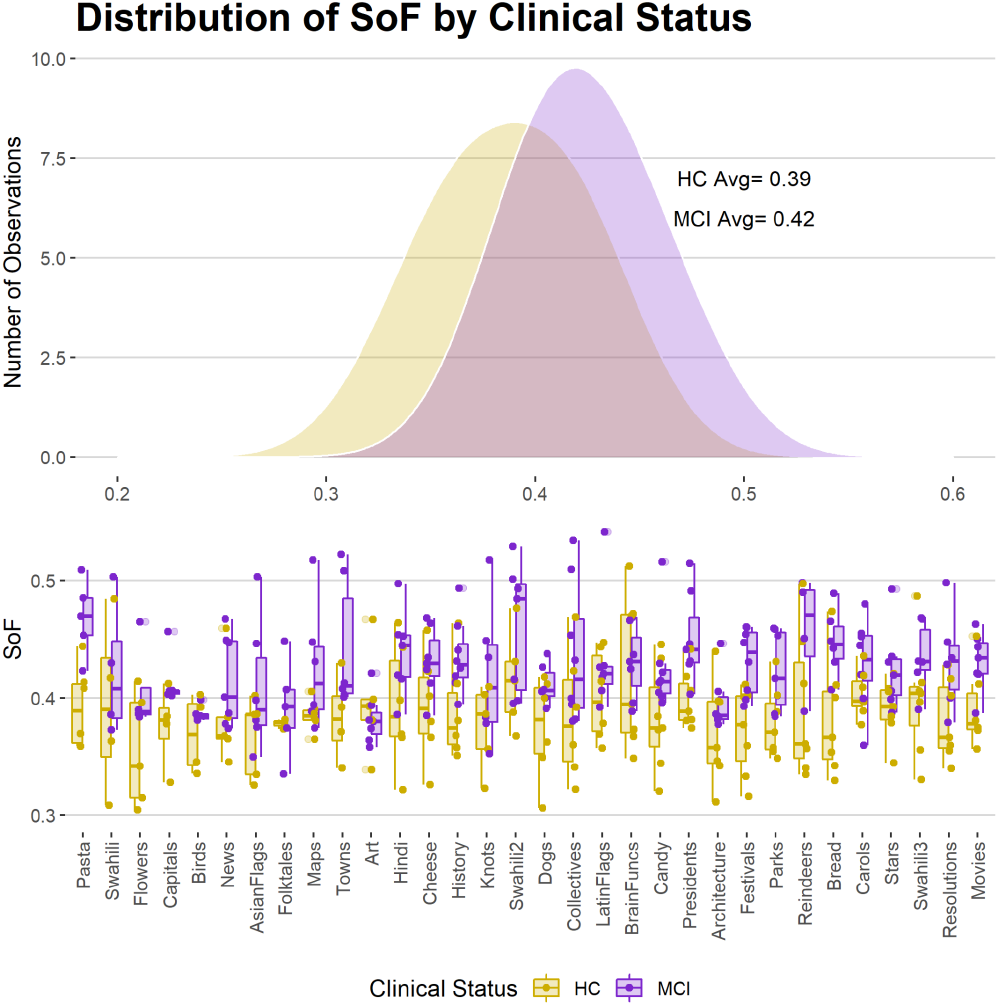
Distribution of *speed of forgetting* by clinical status across all weekly lessons.

### Diagnostic Validity of the *Speed of Forgetting*

To analyze the parameter’s diagnostic potential, its classification accuracy was plotted with a receiver operating characteristic (ROC) curve. The ROC curve assesses the sensitivity (true positive rate) and specificity (true negative rate) of a classifier for varying thresholds of the speed of forgetting parameter. The overall accuracy of the classifier is then measured as the Area Under the Curve (AUC) of the sensitivity and specificity obtained for different thresholds. First, we examined the ROC curve for a single 8-minute session of data, that is, the probability of correctly identifying group members (MCI or controls) by the *speed of forgetting* value of a single test (Fig 5A). The model proved to be highly diagnostic, with an AUC = 0.786 (that, is, a classification accuracy of 78.6%). Then, we examined the ROC curve for a classifier built on the average speed *of forgetting* of an individual, computed across all sessions. As expected, the classifier showed an improved accuracy of 83.6% (Fig 5B). Note that, because of the high test-retest reliability of the *speed of forgetting*, classification using a single session is almost as accurate as when averaging over 30+ sessions.

**Figure 5.**
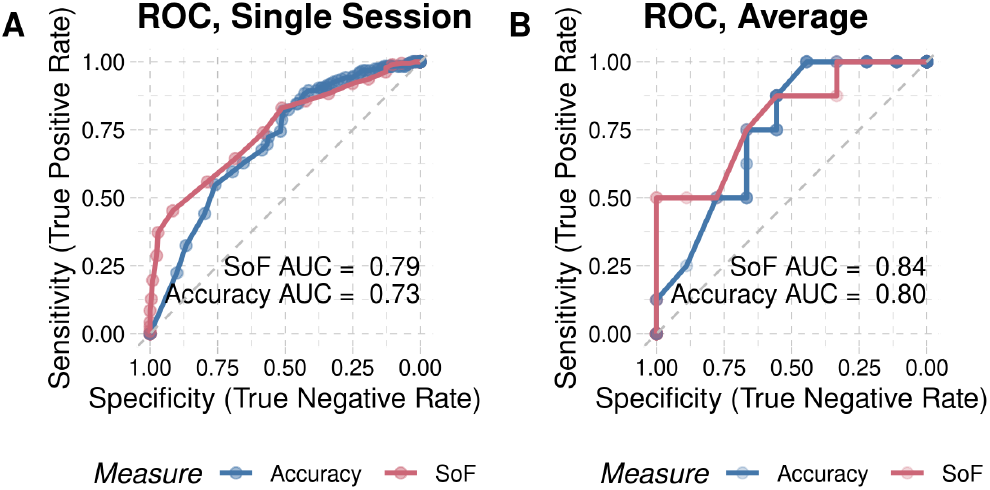
ROC classification performance for the *speed of forgetting* (red) and for response accuracy (blue) from (A) a single session, and (B) averages across sessions.

We also investigated whether the *speed of forgetting* value provided additional validity over traditional behavioral measures, such as response accuracy. Note that, accuracy and response times are collected from the AFLS, which already uses the *speed of forgetting* to determine the best moment at which a study pair is presented. Thus, the validity of these traditional behavioral measures is, in fact, inflated because they benefit indirectly from the study items being driven by the *speed of forgetting*. Despite this, a classifier based on response accuracy alone had a lower AUC than a classifier built on *speed of forgetting*, both for single sessions and for aggregated data (Figures 5A and 5B, blue lines). Specifically, in both cases, an SoF-based classifier showed greater sensitivity than an accuracy-based one even at high levels of specificity (lower left corners of the ROC plots). That is, SoF allows for a greater ability to detect mild memory impairments while still avoiding false positives.

Finally, a logistic curve was generated to model the probability of the binary diagnosis outcomes (Fig 6). To generate the model, individual *speed of forgetting* values for each lesson were binned in increments of 0.01, and the probability of MCI diagnosis was computed as the proportion of individuals with an MCI diagnosis for each bin. The logistic model showed a strong fit to the data (Cragg and Uhler’s pseudo *R*^2^ = 0.26). The inflection point of the curve (that is, the point at which the probability of an MCI diagnosis is > 50%) was found to be at a value of *α* = 0.40. Additionally, the curve revealed that the probability of an MCI diagnosis occurring increased steadily as the *α* value increased, approaching a maximum probability of 1.0 at *α* = 0.52. Overall, the logistic curve provided a clear visualization of the relationship between the predictor *speed of forgetting* and a diagnosis of MCI.

**Figure 6.**
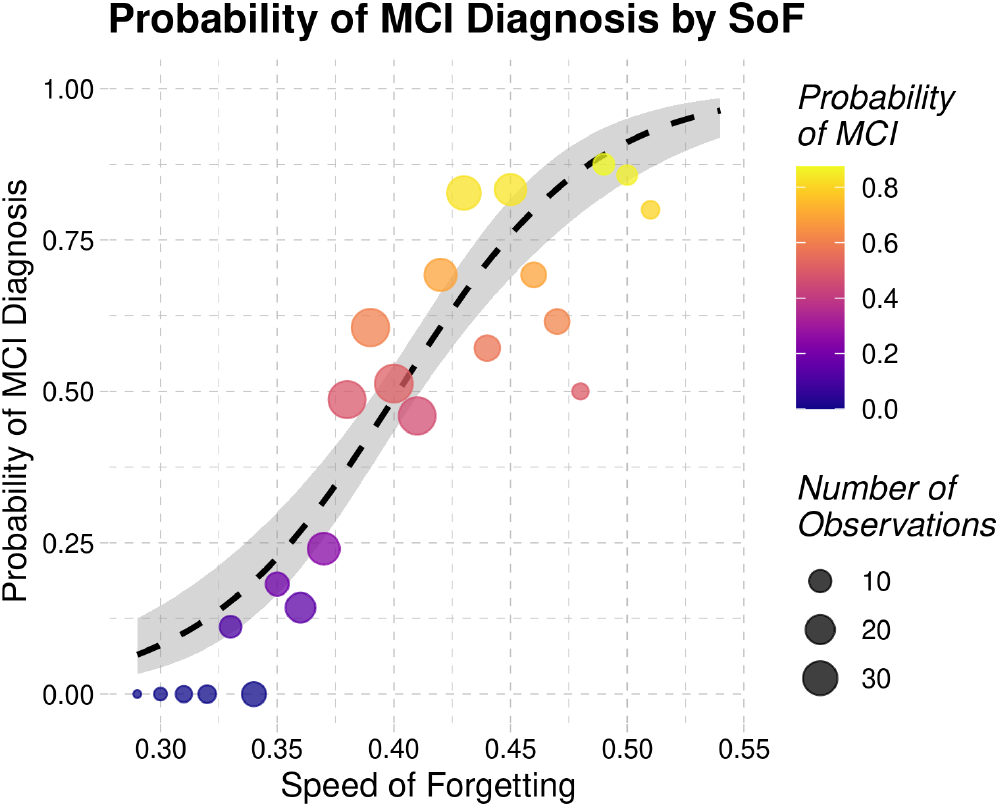
Probability of MCI by *speed of forgetting*.

### Memory Assessment Temporal Trends

As memory function worsens in MCI patients, *speeds of forgetting* should steadily increase over time. While participants were only halfway through the year-long experiment, the subtle changes in the longitudinal trajectory of MCI patients can already be seen (Fig 7). The effect of time was captured using a mixed-effects linear model that included the week number as the main factor and the weekly topic as a random effect (to account for differences in difficulty across topics). In other words, forgetting grew significantly in our sample over time, by approximately 0.15% per week. However, no significant interaction between time and group existed. The analysis uncovered a significant effect of the week (β = 0.0005, *t* = 2.06, *p* = 0.04). Further analysis showed that this effect was driven by the MCI group alone; separate linear models uncovered a significant effect of the week for the MCI (β = 0.0006, *t* = 2.02, *p* = 0.04) but not for healthy controls (β = 0.0003, *t* = 1.13, *p* > 0.26). Thus, although the *speed of forgetting* grew almost twice as fast as in controls, a significant difference in the rate of growth could not yet be detected within our limited sample and time window.

**Figure 7.**
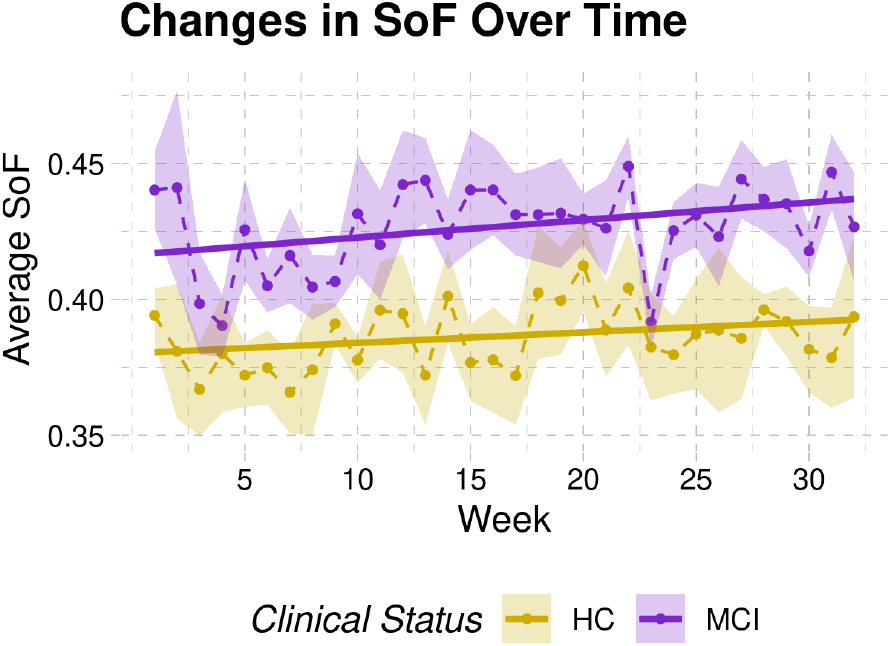
Time course of forgetting.

### Differences Between Materials

As noted above, this longitudinal study also offered the opportunity to examine the effects of different types of materials (verbal vs. visual) on memory function. To assess if the type of stimulus material presented affected lesson difficulty, *speed of forgetting* for verbal stimuli and visual stimuli lessons were compared. Healthy controls had similar *speeds of forgetting* averages (verbal *α* = 0.390; visual *α* = 0.383), but MCIs had a higher *speed of forgetting* for verbal stimuli (verbal *α* = 0.430; visual *α* = 0.416) (Fig 8). Upon further investigation, it was found that the main stimulus features contributing to differences in verbal and visual materials for MCIs were non-English language and numeracy (both of which were primarily in verbal materials). But, it is possible that the visual stimuli also simply provided more features to aid in memorization.

**Figure 8.**
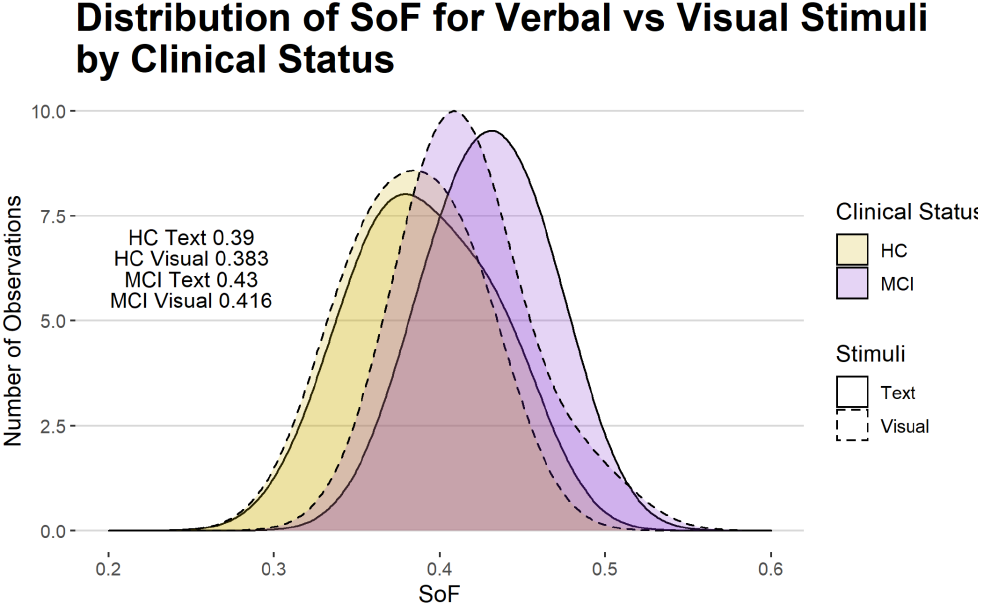
*Speed of forgetting* by clinical status and verbal vs visual stimuli.

### MCI Subtype

To explore the model’s ability to parse out the nuances in cognitive deficits, we categorized MCI groups subtype. We further demonstrated that it was possible to use *speed of forgetting* to accurately distinguish MCI subtypes, including amnestic single and multiple domains (aMCI S, aMCI M) and nonamnestic MCI (naMCI). The aMCI subtype is characterized by a specific memory impairment, while the naMCI subtype is characterized by a more general cognitive decline (Fig 9). The results revealed that the cognitive profile of the naMCI participant more closely resembled that of the healthy control group. This observation is in line with the fact that naMCI is characterized by a cognitive decline in domains other than memory, such as executive function (e.g. speed of processing, problem-solving, set-shifting, inhibition). Therefore, it is expected that the data would be more comparable to the control group as there is no memory loss present in naMCI. However, the model showed limitations in accuracy for the single dementia patient, likely due to differences in response time and reduced facts retained, requiring further refinement for this population.

**Figure 9.**
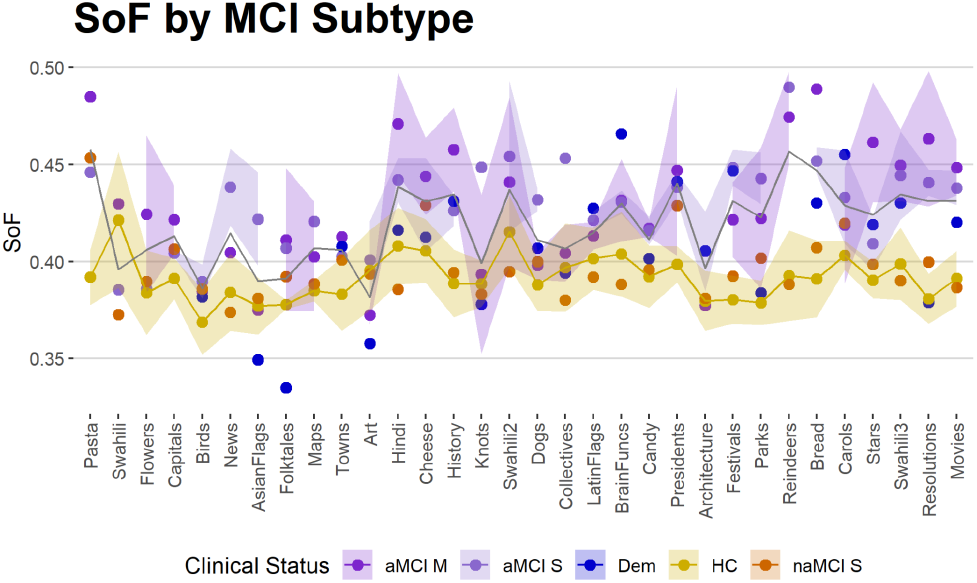
*Speed of forgetting* across lessons by subtype.

## Discussion

Our study presents a novel approach for tracking and diagnosing mild memory impairments using the *speed of forgetting* model parameter from a computational cognitive model. While this model was originally developed for student fact learning, it has never been explicitly used in clinical populations. Here, the *speed of forgetting* was found to be normally distributed, with higher means and ranges in MCI, and showed high diagnostic validity with test-retest reliability. The model also revealed differences in MCI verbal and visual memory, MCI subtypes, and subtle declines over time. Verbal vs visual differences could pinpoint left hemisphere involvement. AD is often asymmetric, yet clinics primarily use verbal memory to diagnose MCI. This model could reduce the ascertainment bias towards right-lateralized MCI. In the example of aMCI vs naMCI, *speed of forgetting* proved to be a purer assessment of memory impairment by avoiding confounds with retrieval strategy and executive function- a common problem in the clinical assessment of mild memory impairment. The ability to track memory over time is of particular importance as early detection of MCI is likely to be crucial in therapies to delay AD and related conditions; and the brief, user-friendly online format makes passive data assessments remarkably convenient. Our data are the first to demonstrate this diagnostic potential, moreover, the repeatability/stability of the measure makes it a good candidate to test the efficacy of interventions like neuromodulation or cognitive enhancers.

### Limitations

As with any new method, there are several limitations that must be considered. First, the sample size used in this study was relatively small, impacting the generalizability of the results to larger populations. We are currently accumulating more data with larger and more diverse sample sizes to confirm the validity of this model in diagnosing memory impairments in different demographic groups.

Second, *speed of forgetting*, in essence, measures the speed of *passive* forgetting, and it is not yet known exactly what that speed reflects. Passive forgetting, i.e. the loss of information over time due to the passage of time rather than a deliberate attempt to forget, could be due to errors in encoding, retrieval, or simply changes in the way that the brain processes information. More research is needed to determine which of these passive mechanisms is most closely linked to this parameter.

Third, this study has only examined recognition memory, not recall memory. Recognition memory refers to the ability to identify previously learned information, while recall memory refers to the ability to retrieve information from memory without any cues or prompts. Research has shown that recognition and recall memory can be affected differently in individuals with MCI (Bennett et al., 2006). For example, individuals with MCI may have relatively preserved recognition memory but impaired recall memory, or vice versa. Therefore, the current measures of recognition memory may not provide a complete picture of the individual’s memory abilities. But, data from recall trials are currently being investigated.

Finally, the computational model showed limitations in accurately diagnosing memory impairment in individuals with *dementia*, likely due to significant differences in response times and the reduced number of facts the participants were able to retain, highlighting the need for further refinement of the model for this population.

### Broader Impacts

This model holds tremendous promise for improving our understanding of memory problems in other psychiatric disorders. Memory impairment is a hallmark of dementia and AD but is also present in other disorders such as schizophrenia, depression, bipolar, PTSD, and ADHD. Each disorder affects memory differently: schizophrenia affects working memory, depression and bipolar impact recall of personal memories, PTSD reduces encoding and recall, and ADHD affects short-term memory and working memory. These examples highlight the diverse and complex ways that memory impairment can manifest, making it a critical area of research.

#### Disciplinary Diversity & Integration

In conclusion, this study represents an important step forward in the burgeoning interdisciplinary field of *computational psychiatry*. Computational psychiatry aims to provide an explanatory bridge between altered cognitive function and its underlying neurobiological mechanisms by means of computational models. We aim to integrate the model with neuroimaging data to discover the underlying neural mechanisms behind memory decline in aging and disease. Clinical applications are not commonly presented in Cognitive Science, and translational applications of cognitive models do not typically include health care and medical diagnosis. This integrative approach can provide valuable insights into the cognitive changes that occur in these conditions and may help to identify new ways to mitigate or prevent these changes.

## Data Availability

All data produced are available online on GitHub, an online data and code repository

